# P5 promoter-mediated incorporation explains REP/CAP manufacturing contaminants in patient liver after rAAV gene therapy

**DOI:** 10.64898/2026.09.10.26362787

**Authors:** Mark A. Brimble, Shaoyuan Tan, Sarah Buddle, Li-An K. Brown, Judith Breuer, Jeremy Chase Crawford

## Abstract

Sequencing of liver tissue from a patient treated with the rAAV gene therapy Zolgensma for spinal muscular atrophy recently revealed contaminating plasmid sequences derived from rAAV manufacturing within the patient’s hepatocytes. In particular, REP/CAP-derived sequences were remarkably abundant, corresponding to 0.5-1% of the therapeutic transgene. We hypothesized that these contaminants originated through Rep-mediated incorporation initiated at the AAV P5 promoter. Through reanalysis of the sequencing data, we inferred that an intact P5 promoter had been placed directly downstream of the rAAV capsid gene in the manufacturing plasmid. *De novo* assembly revealed a contiguous contaminant sequence spanning the rAAV REP and CAP genes and terminating within P5 at the Rep nicking site, immediately downstream of the Rep-binding element (RBE). This analysis also revealed a distinct vector-plasmid backbone contig consistent with reverse packaging. Among partially aligned REP/CAP reads, nearly 17% were linked to rAAV ITR-derived sequence at heterogeneous junctions. Long-read data independently identified the P5 promoter as the most frequent recombination breakpoint region. Together, these findings identify a defined, avoidable mechanism by which REP/CAP manufacturing contaminants arise. Positioning of the P5 promoter within the manufacturing plasmid is therefore a modifiable determinant of rAAV product purity and the transfer of rAAV DNA contaminants to recipient patients.

## Introduction

Recombinant adeno-associated virus (rAAV) is the most widely used vector platform for *in vivo* gene therapy; thousands of patients have received rAAV-based therapies,^1^ and there are now ten rAAV drug products approved by the FDA for a range of monogenic disorders.^2,3^ However, serious adverse events, in some cases with fatal outcomes, have been associated with rAAV administration.^4–8^ Given the heterogeneity of dose, administration route, and disease context, identifying the factors that contribute to these adverse events is increasingly important as the treated population expands. For Spinal Muscular Atrophy (SMA), the FDA-approved rAAV-based drug Zolgensma (onasemnogene abeparvovec-xioi) has dramatically improved event-free survival and motor function for treated neonatal patients.^9^ Yet, rare life-threatening complications, including Hemophagocytic Lymphohistiocytosis and acute fatal liver failure, have been reported following the administration of Zolgensma.^10^ Understanding how patient-specific factors, design of the gene therapy, and ultimate composition of the drug product contribute to risk is essential for interpreting and mitigating adverse events observed in real-world clinical contexts.

Residual manufacturing DNA packaged within vector particles is a recognized impurity in rAAV gene therapies, raising questions about the abundance, origin, persistence, and biological consequences of these contaminants following patient treatment. The dominant mechanisms by which plasmid-derived DNA is incorporated and encapsidated into rAAV particles have been elucidated in recent years.^11–15^ In particular, sequences adjacent to AAV Rep-protein-binding elements (RBEs) are preferentially incorporated and can exceed 1% of total vector DNA content.^16^ In standard rAAV production workflows, these RBEs are typically found within the ITRs flanking the vector genome and in the native P5 promoter driving large isoform Rep protein expression.

Contaminating plasmid sequences derived from rAAV manufacturing were recently identified in the liver of a child who developed hepatitis after treatment for SMA with Zolgensma.^17^ Manufacturing-derived AAV (REP/CAP) sequences were detected in ~5% of hepatocytes and found at levels inconsistent with stochastic incorporation of plasmid DNA during vector manufacturing. These observations raise the question of whether the contaminants originated through a defined packaging mechanism rather than random encapsidation. Here, through mechanistic reanalysis of patient sequencing data, we conclude that the high levels of REP/CAP contaminants arose via Rep-mediated incorporation initiated at the P5 promoter during rAAV manufacturing. Importantly, because their incorporation results directly from P5 positioning within the manufacturing plasmid, these contaminants are avoidable. Although P5-associated sequences have been characterized in rAAV preparations and preclinical models, our findings demonstrate that these manufacturing-derived contaminants are transferred to patients and can be detected at high abundance in human tissue following rAAV gene therapy.

## Results

During rAAV production, P5-mediated encapsidation of contaminating plasmid DNA occurs unidirectionally upstream of P5 and independently of the packaging of the ITR-flanked vector genome (Figure 1a).^11^ We therefore sought to determine whether the abundant REP/CAP sequences identified in patient liver^17^ arose from this mechanism. Because the proprietary plasmid production sequences used for Zolgensma are not publicly available, we reasoned that the representative mapping references used by Buddle et al.^17^ cannot be considered ground truth and may differ from the actual manufacturing constructs. We therefore conducted *de novo* assembly of the human-filtered dataset, which resolved two multi-kilobase contigs (Figure 1b, Supplementary Figure 1). Node 1 contained the complete REP and CAP coding sequences derived from the packaging plasmid, whereas Node 2 contained contiguous plasmid backbone sequence. Because sequences directly adjacent to ITRs are preferentially incorporated as contaminants into rAAV particles,^13,18,19^ the high abundance of Node 2 (~3% of the vector genome; Supplementary Table 1) is consistent with the previously proposed mechanism of reverse packaging of undersized (<5kb) plasmid backbone.^12^

**Figure 1.**
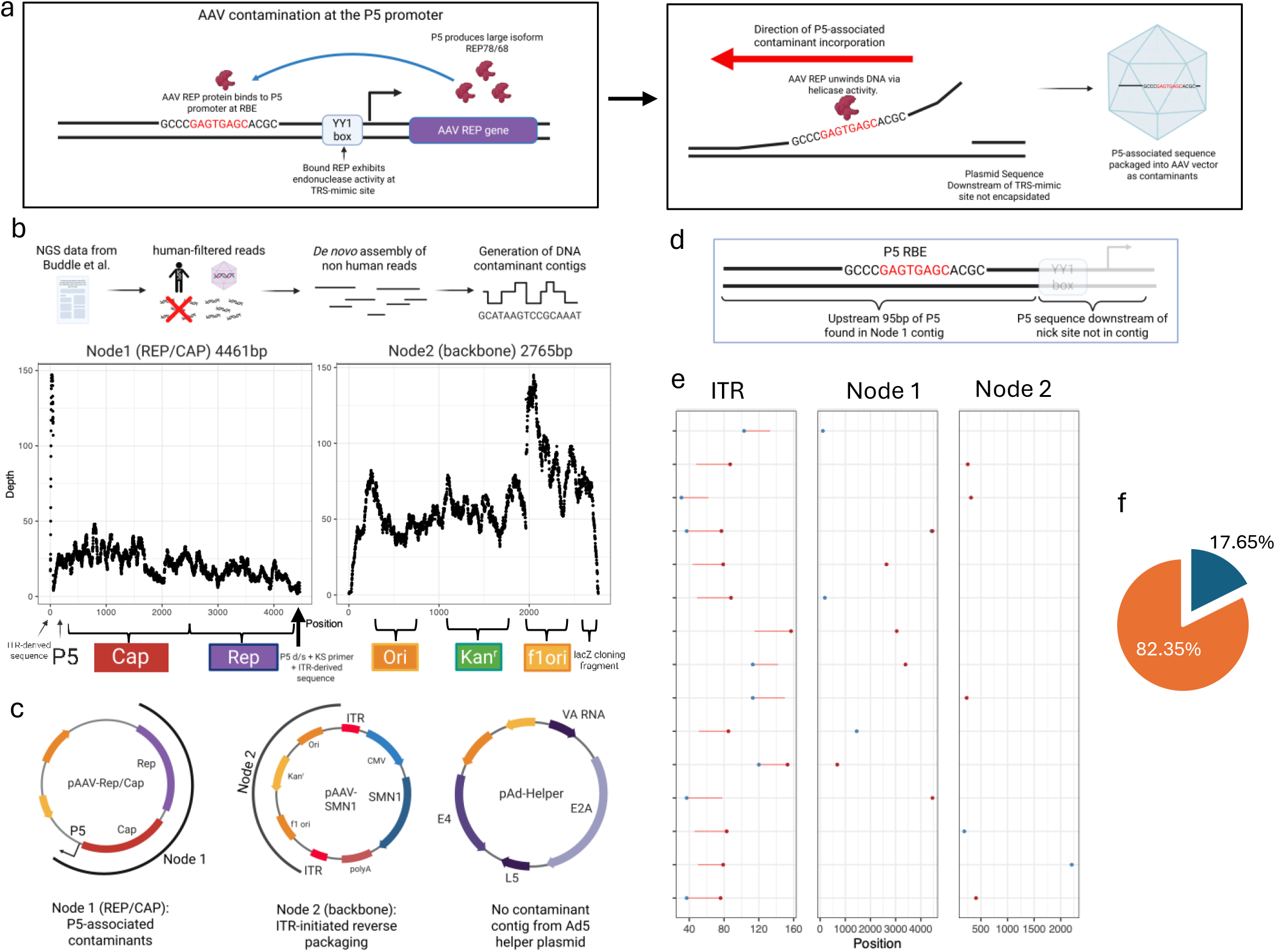
Identification of P5 as the origin of REP/CAP DNA contaminants within the liver biopsy of a Zolgensma-treated patient. (a) Schematic depicting mechanism of P5-associated contamination in rAAV (top). The P5 promoter drives expression of large isoform rAAV replication proteins (REP78/68). These proteins bind (via negative autoregulation) back to the P5 promoter to repress transcription. Bound Rep is able to nick via endonuclease activity within the YY1 transcription factor binding site (YY1 bs). Rep exhibits helicase activity (bottom) and is able to extrude plasmid sequence upstream from the nick site, which is then actively packaged into the preformed rAAV capsid and observed at high levels within the rAAV drug product. (b) *De novo* assembly of rAAV contamination contigs from human-filtered reads obtained from liver biopsy. (Left) Node 1 contig contains the rAAV replication (REP) and capsid (CAP) genes linked to P5 and ITR-derived sequence. (Right) Node 2 contig contains a contiguous region of contaminant plasmid backbone elements. (c)Schematic illustrating likely node origins within the rAAV manufacturing plasmids used to produce Zolgensma. (Top) The rAAV REP/CAP packaging plasmid from which Node 1 is inferred to originate. (Middle) The ITR-flanked vector plasmid, from which Node 2 is inferred to derive by reverse packaging of plasmid backbone sequence. (Bottom) A representative adenovirus helper plasmid, which does not appear to contribute substantially to the abundant, actively incorporated DNA contaminants identified in this study. (d)The assembled Node 1 contig terminates at the rAAV P5 Rep nicking site. Upstream plasmid sequence is retained, whereas no downstream sequence is observed, consistent with the mechanism of Rep-mediated, P5-associated incorporation. (e)Soft-clipped Illumina reads support heterogeneous linkage between rAAV ITR-derived sequence and the assembled contaminant contigs. The clipped portion of each read maps to the rAAV ITR (left), whereas the aligned portion maps to Node 1 (REP/CAP; middle) or Node 2 (backbone; right). The broad distribution of junction positions across contigs is suggestive of heterogeneous concatemer formation. Blue dot, left-end soft-clipped reads. Red dot, right-end soft-clipped reads. (f)Pie chart showing percentage of REP/CAP contig-aligned, soft-clipped reads that also aligned to AAV ITR sequence (blue) or any other sequence (orange).

Examination of the assembled REP/CAP contig (Node 1) revealed a native P5 promoter positioned directly downstream of the capsid gene. This configuration, historically theorized to enhance production efficiency through the promoter enhancer function that P5 effects on the AAV P19 and P40 promoters,^20,21^ is common in manufacturing constructs.^11,14,18^ However, placing the P5 promoter in this position is not required for high-titer AAV manufacturing at clinical scale.^22^ Notably, the assembled contig terminated within P5 without extending into adjacent plasmid sequence (Figure 1d). This breakpoint is located within a few nucleotides of the characterized Rep nicking site in P5.^23^ Considered in conjunction with our recovery of upstream REP/CAP sequence and absence of downstream plasmid sequence, this sequence boundary strongly supports Rep-mediated incorporation initiated at the P5 promoter as the mechanism generating the REP/CAP DNA species observed in this biopsy. The assembled sequence also indicates that only the downstream region of P5, a region approximately 45bp in length and devoid of the RBE, is present directly upstream of the AAV Rep gene. This is clearly plasmid-derived, flanked by a plasmid-backbone-specific KS primer sequence. Surprisingly, both ends of the *de novo* assembled Node 1 contig were joined directly to ITR-derived sequence, despite the absence of ITRs in the REP/CAP manufacturing plasmid. This structure suggests post-transduction recombination between the REP/CAP contaminant and ITR-containing rAAV DNA and is consistent with incorporation of these contaminants into stable concatemers.

These concatemeric structures could account for the plasmid-derived DNA contaminants that exceed the packaging limit of AAV vectors described in the original study.^17^

Since P5-associated contaminants comprise fragments of heterogeneous lengths,^11^ we also examined sequences only partially aligned to the REP/CAP node to ascertain where recombination had occurred. 17% (9/46) of soft-clipped reads spanning Node 1 were linked to AAV ITR-derived sequence at heterogeneous positions across the REP/CAP contig (Figure 1e, f), consistent with recurrent recombination between manufacturing-derived contaminants and ITR-containing rAAV DNA. Recombination between the plasmid backbone sequences in Node 2 and ITR-derived sequence was also detected, predominantly near the ends of the node (Figure 1e).

To determine if long-read sequencing independently supported this apparent recombination, we reanalyzed available nanopore data (ONT) from the same patient. Coverage was limited, with only 16 reads mapping to the REP/CAP contig. Consistent with Buddle et al.^17^, we did not identify a full-length replication-competent AAV genome. However, analysis of sequence breakpoints along the length of Node 1 identified P5 as the most frequent recombination breakpoint region, with P5 breakpoints observed in 5/16 reads (31.25%; Figure 2a). One read contained two independent P5 sequences, indicating that a short P5-initiated contaminant fragment recombined with a second, larger fragment (Figure 2b). Together, these analyses of long-read data independently support P5 as the predominant site of contaminant initiation and recombination. Direct recombination between node sequences and AAV ITR sequence was also identified in the long-read data from individual reads that mapped to both AAV ITR sequence and either of the *de novo-*assembled node references generated from the short-read dataset (Figure 2c). Due to lower sequencing depth, this was detected in only a single instance for each node. Notably, Node 1 contained a largely intact AAV ITR sequence directly linked to the 5’ region of the AAV Rep gene. As with the original analysis, no full-length REP/CAP DNA species were resolved from the data.

**Figure 2.**
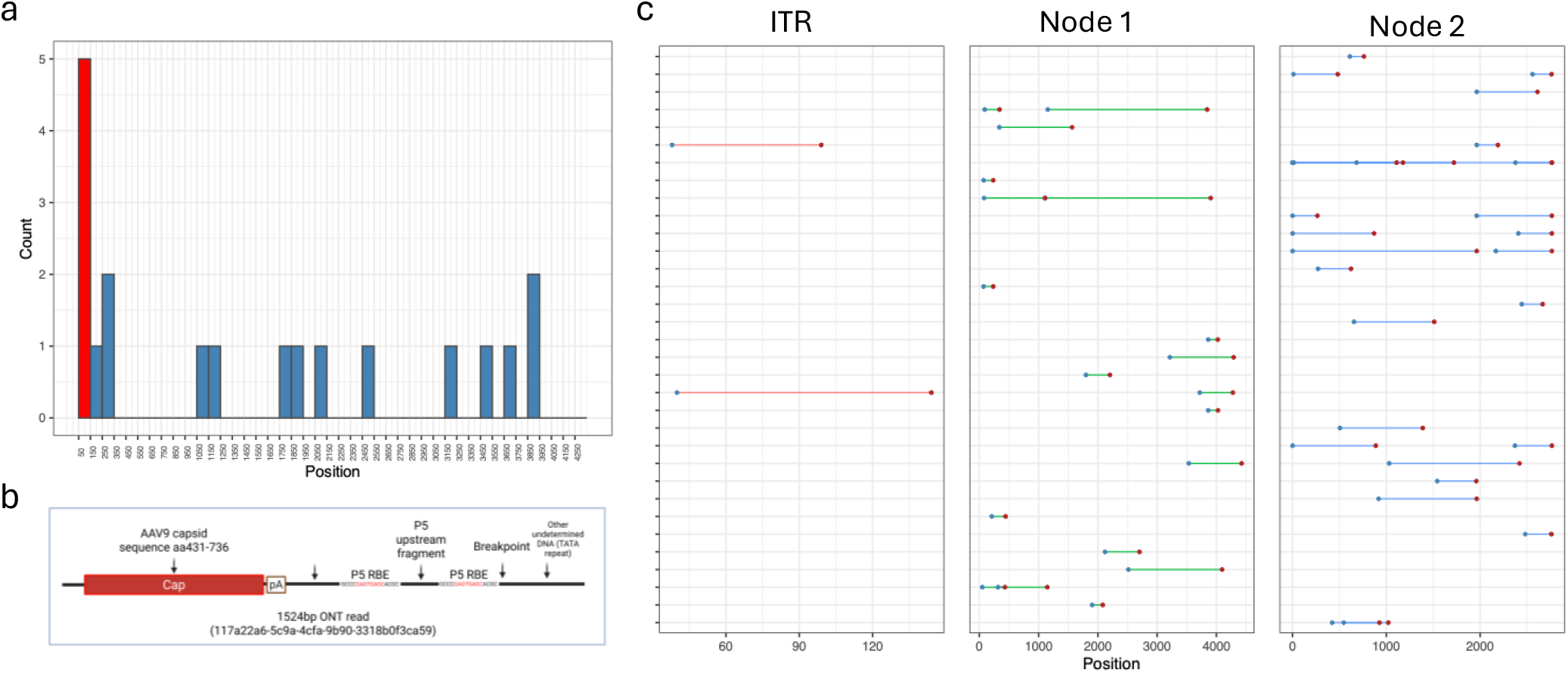
Mapping of ONT sequencing reads to *de novo* assembly identifies the P5 promoter as a preferred recombination breakpoint. (a)Histogram analysis of the number of ONT reads containing recombination breakpoints within a given region of the Node 1 REP/CAP contig. Each bar represents a 100bp region of Node 1. Red bar, P5 promoter region. (b)Schematic of an ONT single-molecule read containing two independent copies of the P5 upstream sequence, indicating recombination of two P5-associated contaminant molecules. (c)ONT long reads mapped to *de novo*-assembled nodes. Line color signifies reference mapping (red = AAV ITR sequence, green = Node 1 REP/CAP contig, blue = Node 2 plasmid backbone contig). Deviations from the mapped node are indicated by colored dots (Blue dot = left-end deviation; Red dot = right-end deviation).

Collectively, these findings support a mechanistic model explaining the high abundance of REP/CAP contaminants identified in patient liver^17^ (Figure 3a). In this model, the P5 promoter positioned immediately downstream of the AAV Capsid gene, together with its AAV RBE and proximal nick site, directly promotes the incorporation of upstream plasmid-derived DNA, which in the case of Zolgensma results in incorporation of the upstream REP/CAP coding sequences as contaminants. This process generates heterogeneous contaminant fragments (up to the rAAV packaging limit of ~5kb), which are packaged into vector particles and subsequently copurified with particles containing the therapeutic vector genome. This model explains both the unexpectedly high abundance of REP/CAP contaminants and their recurrent structural association with rAAV-derived sequences. To determine how prevalent such a configuration is among commonly used plasmids for AAV production, we examined the top 30 most requested AAV capsid plasmids from the plasmid repository Addgene (Figure 3b; Supplementary Table 2). 77% (23/30) of AAV serotype REP/CAP plasmids contained a full-length P5 promoter downstream of the capsid gene and would therefore be predicted to incorporate wildtype AAV genes during production; the remaining 23% (7/30) contained an intact P5 promoter either directly or within a few hundred base pairs upstream of the AAV Rep gene and would instead be susceptible to incorporating plasmid backbone sequences.

**Figure 3.**
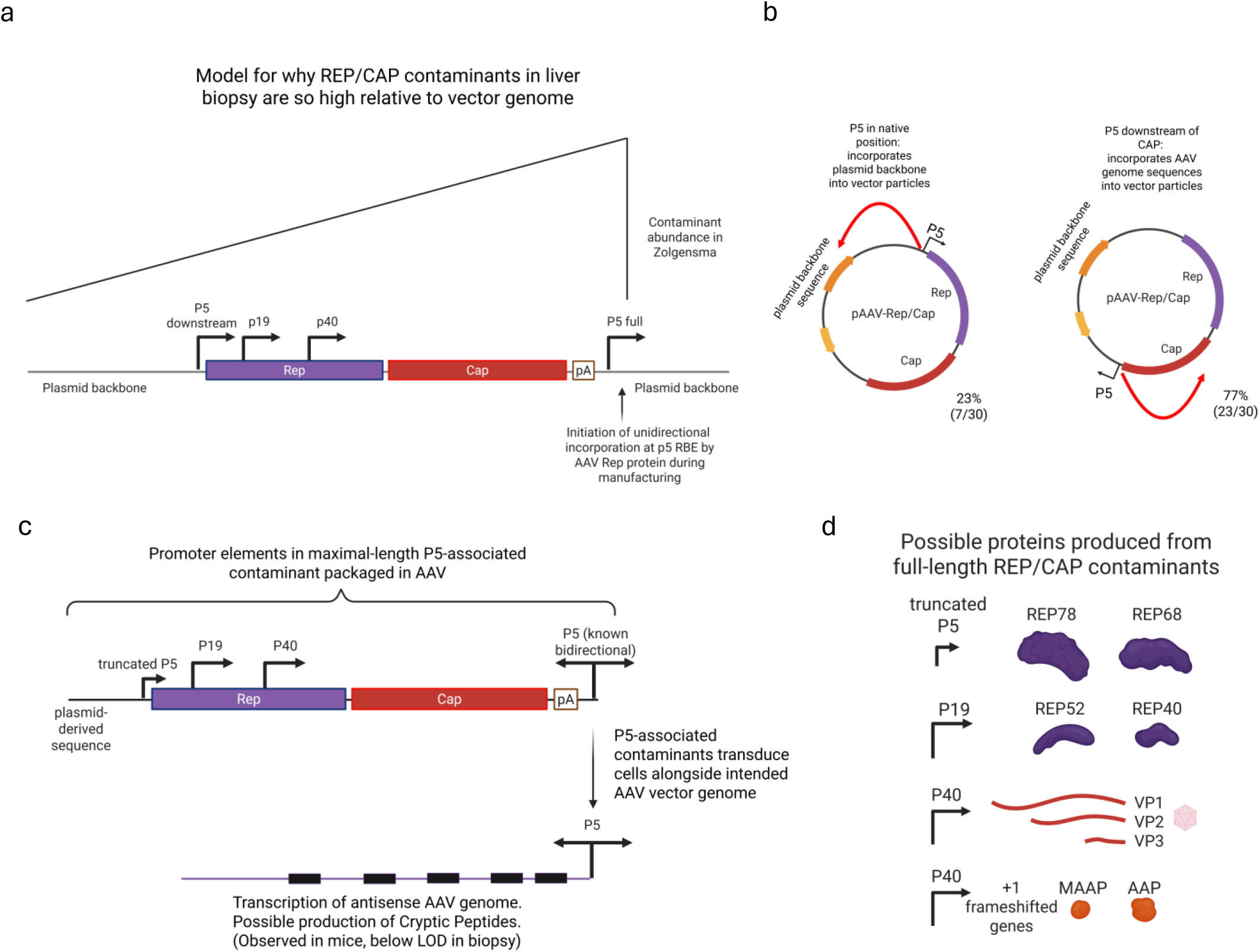
Model of P5-associated contamination and its potential downstream effects. (a)Proposed mechanism for P5-mediated REP/CAP contaminant formation during rAAV manufacturing. Black 90-degree forward arrows indicate canonical transcription direction from rAAV promoter sequences. Due to contaminant initiation at the downstream P5 sequence, and the propensity of these sequences to comprise fragments of heterogeneous lengths, the contaminant abundance of specific sequences in the rAAV genome relates to the relative distance from P5. (b)Frequency of most requested (top 30) AAV serotype plasmids in which P5 is placed in its native position directly upstream of REP (left) or in the configuration inferred for Zolgensma, with P5 placed directly downstream of the capsid gene (right). Red arrows depict the direction and initiation point of these contaminants. (c)Proposed transcriptional potential of full-length REP/CAP contaminants. Bidirectional activity of the retained P5 promoter (black forward and reverse arrow), together with known upstream transcriptional activity of P5-associated contaminants, could drive antisense transcription of the transferred contaminant, possibly yielding cryptic protein products. (d) Retention of the native rAAV promoter architecture (truncated P5, P19, P40) also preserves the potential for expression of wildtype AAV gene products from larger DNA contaminant species transferred to the patient.

## Discussion

In this study, mechanistic reanalysis of patient sequencing data identified P5-mediated incorporation as the origin of highly abundant wildtype AAV REP/CAP manufacturing contaminants recovered from a post-Zolgensma liver biopsy.^17^ By performing *de novo* assembly of the vector contaminants, we established a ground truth reference directly from the patient sequencing data, allowing us to resolve two contiguous plasmid-derived contaminants that were previously obscured. Mapping to this reconstructed reference identified the P5 promoter as a preferential recombination breakpoint. Together, these findings indicate that the high abundance of REP/CAP contaminants results from P5-mediated incorporation, while their sequence content is determined by the nonessential placement of the P5 promoter downstream of the capsid gene in the manufacturing plasmid. These findings raise three key considerations for rAAV manufacturing and product safety. First, placement of P5 directly downstream of the capsid gene creates a mechanism for transferring near- or full-length rAAV genomes that can persist within the transduced cells of the treated individual. Second, the heterogeneous linkage to ITR-sequence we observed provides a pathway through which these contaminants could establish replication-competent wildtype-like AAV genomes. Lastly, because this placement of P5 is unnecessary for efficient rAAV manufacturing, these findings identify a tractable opportunity to reduce this class of contaminants in rAAV therapeutics.

While our analysis identified P5 as the contaminant sequence boundary and a recurrent recombination breakpoint in biopsy sequencing data, prior published work provides the mechanistic framework for concluding that P5 serves as the replication origin driving the formation of these contaminants. The ability of P5 to serve as an alternative replication origin in the absence of an AAV ITR has been characterized previously,^23^ and in the context of recombinant AAV it is known that P5 can incorporate DNA up to the 5kb packaging limit during manufacturing, and that such incorporation during manufacturing occurs independently of the presence of an ITR-flanked vector genome.^11^ Further supporting this mechanism, it has been shown that direct modification of the Rep nicking site in P5 can dramatically reduce the abundance of these P5-associated contaminants.^24^

The inferred structure of these contaminants also has important biological implications. P5 exhibits bidirectional promoter activity,^11^ and P5-associated contaminants have been shown to be transcriptionally active following rAAV transduction in cell culture and the livers of rAAV-infected mice.^11^ Protein translated from P5-associated contaminants is even sufficient to elicit *de novo* T cell responses.^11,25^ The RNAseq read-depth in Buddle et al. ^17^ was insufficient to determine if REP/CAP contaminants were transcribed in this biopsy. Additional studies with higher sequencing depths are required to determine whether the transcriptional activity observed experimentally also occurs following clinical rAAV gene therapy. With the manufacturing configuration identified from this biopsy, transcription initiated from these P5-associated contaminants would occur in the reverse direction relative to the rAAV genome and could produce cryptic peptides if translated. However, because these contaminants retain the native arrangement of the downstream P5 sequence together with internal AAV promoters (P19 and P40), expression of large-form REP78/68, short-form REP52/40, the AAV Capsid, and the other associated proteins (AAP/MAAP) may also be possible from these REP/CAP contaminants (Figure 3c, d).

An AAV vector manufacturing design in which P5 is placed downstream of the capsid gene remains widely used in the field. Its prevalence is likely due to the field-wide dissemination of this early plasmid design that could produce high-yield AAV vectors irrespective of transgene.^20^ The ability of the P5 promoter to incorporate upstream DNA as contaminants was elucidated two decades after this design was constructed.^11,14,18^ The P5 promoter has been kept in rAAV manufacturing systems in part because the Rep protein needs to be tightly regulated for efficient production.^26^ It should be noted that even if the native P5 promoter is positioned upstream of Rep, it will incorporate plasmid backbone sequences into the rAAV particles. It is arguable that this would be preferable to incorporating the wildtype Rep and Cap sequences, but either configuration will result in the transfer of DNA contaminants at abundances often exceeding 1% of the total rAAV DNA content.^16^ Production strategies that reduce the presence of P5-associated contaminants by orders of magnitude while retaining high AAV yields have been developed,^11,24^ as have systems that do not rely on the P5 promoter to facilitate Rep gene expression.^27^ For instance, baculovirus-based AAV production in insect cells typically uses a heterologous promoter such as polyhedrin or ΔIE1 to drive expression of the AAV Rep gene.^28,29^

Since we were analyzing data from tissue and not from the drug product itself, it is possible that the levels of REP/CAP contaminants in the Zolgensma drug product are not as high as those observed in the biopsy. This could occur if these AAV-genome-derived contaminants had a greater ability to persist than therapeutic cassette DNA, or if the REP/CAP DNA had undergone replication within the patient. Public reporting of viral preparation integrity from this and other clinically approved rAAV drug products would greatly aid efforts to understand the root causes of, and ultimately prevent, toxicities associated with rAAV gene therapy.

The clinical success of Zolgensma highlights the promise of gene therapy. However, several high-profile instances of toxicity after rAAV gene therapy have occurred within the field over the last eighteen months, often with unexplained root causes.^4–7^ Assuming equivalent DNA persistence and no replication of the wildtype AAV genes within the Zolgensma-treated patient analyzed here, the quantity of plasmid-derived contaminants delivered to this patient would be ~4.3% of the transgene (Supplementary Table 1); this equates to 4.3 × 10^12^ vg/kg when accounting for the dose of Zolgensma, with an estimated REP/CAP contaminant dose of between 5 × 10^11^-1 × 10^12^ vg/kg. By comparison, this would represent a higher delivered quantity of Rep and Cap gene DNA than the therapeutic FIX vector genome administered in the first successful liver-directed gene therapy trial for hemophilia B.^30^ As our understanding of vector-derived impurities evolves, so should the standards used to evaluate and minimize them. Placement of the native, intact AAV P5 promoter immediately downstream of the capsid gene represents a legacy manufacturing design that contributes to the risk profile of rAAV vectors via abundant transfer of wildtype AAV genes to patients. This warrants reevaluation in gene therapy products destined for the clinic.

## Materials and Methods

Sequence data were filtered to remove human reads before transfer to St. Jude Children’s Research Hospital for analysis under the terms of a Material Transfer Agreement and following a St. Jude IRB determination of Non-Human Subjects Research (RB 26-2354). Illumina reads were filtered using a metagenomics preprocessing pipeline utilizing the files generated after human sequence removal using BLAST but before rRNA filtering. Code is available at https://github.com/smorfopoulou/clinical_metagenomics. ONT reads were filtered by aligning to human genome GRCh38.p14 with minimap2 and extracting the non-aligned reads with samtools.

Quality-filtered paired-end Illumina reads were assembled *de novo* using SPAdes. Two contigs (NODE_1 and NODE_2) from the assembly were compiled into a multi-fasta reference for downstream mapping. Illumina reads were aligned to this reference using BWA-MEM, and per-base depth of coverage was calculated with samtools. ONT long reads were aligned to the same reference using minimap2. Soft-clipped reads were identified from the alignments using samtools, and clip characteristics were extracted with a custom Python script. To assess the relationship between soft-clipped reads and inverted terminal repeat (ITR) regions, soft-clipped reads were extracted and re-mapped against an AAV2 ITR reference sequence. All downstream analyses were visualized using custom R scripts. For assessment of ONT breakpoints, alignments were made between nucleotides 50 and 4,424bp to exclude the ITR sequences that would not have been in the original REP/CAP manufacturing plasmid.

## Data Availability Statement

The human-filtered read dataset used in this study is available upon request from Dr. Judith Breuer following the fulfillment of a data transfer agreement.

## Author Contributions

Authors Brimble, Tan, and Crawford requested data from the original study authors to examine this and collaborated with authors Buddle, Brown, and Breuer to interpret this contemporaneous reanalysis of the data.

M.A.B. - Conceptualized, Methodology, Investigation, interpreted data, Writing - Original Draft, Project administration

S.T. – Methodology, Software, Formal Analysis, Writing - Original Draft

S.B. – Resources, Writing - Review C Editing

L.B. – Resources, Writing - Review C Editing

J.B. – Resources, Writing - Review C Editing

J.C.C. – Methodology, Resources, Supervision, Project administration, Writing - Review C Editing

